# Safety and immunogenicity of INO-4800 DNA vaccine against SARS-CoV-2: a preliminary report of a randomized, blinded, placebo-controlled, Phase 2 clinical trial in adults at high risk of viral exposure

**DOI:** 10.1101/2021.05.07.21256652

**Authors:** Mammen P. Mammen, Pablo Tebas, Joseph Agnes, Mary Giffear, Kimberly A. Kraynyak, Elliott Blackwood, Dinah Amante, Emma L. Reuschel, Mansi Purwar, Aaron Christensen-Quick, Nieman Liu, Viviane M. Andrade, Julie Carter, Gabriella Garufi, Malissa C. Diehl, Albert Sylvester, Matthew P. Morrow, Patrick Pezzoli, Abhijeet J. Kulkarni, Faraz I. Zaidi, Drew Frase, Kevin Liaw, Hedieh Badie, Keiko O. Simon, Trevor R.F. Smith, Stephanie Ramos, Robert Spitz, Robert J. Juba, Jessica Lee, Michael Dallas, Ami Shah Brown, Jacqueline E. Shea, J. Joseph Kim, David B. Weiner, Kate E. Broderick, Jean D. Boyer, Laurent M. Humeau

## Abstract

**Background:** Vaccines against SARS-CoV-2 are still urgently needed as only 5% of the global population has been vaccinated. Here we report the safety and immunogenicity of a DNA vaccine (INO-4800) targeting the full-length Spike antigen of SARS-CoV-2 when given to adults at high-risk of exposure.

**Methods:** INO-4800 was evaluated in 401 participants randomized at a 3:3:1:1 ratio to receive either INO-4800 (1 mg or 2 mg dose) or placebo (1 or 2 injections) intradermally (ID) followed by electroporation (EP) using CELLECTRA® 2000 at Days 0 and 28. ClinicalTrials.gov Identifier: NCT04642638

**Findings:** The majority of adverse events (AEs) were of Grade 1 and 2 in severity and did not appear to increase in frequency with the second dose. The number of participants experiencing each of the most common AEs did not differ appreciably between the two dosing groups. The geometric mean fold rise (GMFR) of binding and neutralizing antibody levels were statistically significantly greater in the 2.0 mg dose group versus the 1.0 mg dose group. The T cell immune responses measured by the ELISpot assay were also higher in the 2.0 mg dose group compared to the 1.0 mg dose group.

**Interpretation:** INO-4800 at both the 1.0 mg and 2.0 mg doses when administered in a 2-dose regimen appeared to be safe and well-tolerated in all adult ages. However, the comparative immunogenicity analysis favored selection of INO-4800 2.0 mg dose for advancement into a Phase 3 efficacy evaluation.

**Funding:** The trial was funded by the Department of Defense Joint Program Executive Office for Chemical, Biological, Radiological and Nuclear Defense, (JPEO-CBRND) in coordination with the Office of the Assistant Secretary of Defense for Health Affairs (OASD(HA)) and the Defense Health Agency.

**Research in context:** INO-4800 is among several vaccines being tested against SARS-CoV-2, the virus that causes COVID-19 with the goal of inducing a protective immune response. The DNA vaccine, INO-4800, administered by ID injection followed by electroporation (EP) using the CELLECTRA^®^ 2000 device, induces a balanced immune response that includes engagement of both T cells and B ^1-5^.

**Added value of this study:** This is the first report of a randomized, blinded, placebo-controlled clinical trial of INO-4800, a DNA vaccine targeting the SARS-CoV-2 Spike antigen delivered ID followed by EP, in adults at high risk of SARS-CoV-2 exposure.

## Introduction

SARS-CoV-2, the causative virus of COVID-19, continues to be a major threat to worldwide health and economic productivity. As of April 2021, there have been over 32 million SARS-CoV-2 cases and 575,000 deaths in the United States^6^. Most infected individuals are either asymptomatic or have mild to moderate disease characterized by symptoms such as cough, fever, and mild pneumonia^7^. Previous infection data indicated that 19 percent of infections result in severe, potentially life-threating disease characterized by shortness of breath, rapid respiratory rates, lung infiltrates, and oxygen desaturation <94%^6,8,9^. Due to the rapid viral spread, it is imperative that safe and effective vaccines against SARS-CoV-2 be immediately developed for ease of worldwide deployment to end the pandemic.

Despite the authorization of three vaccines for emergency use in the United States^11-13^ and the release of several more vaccines worldwide^13-16^, SARS-CoV-2 infection continues to outpace the administration of vaccines in several countries across the globe demonstrating that the current vaccine supply is far from sufficient. In India alone, there were 6 million new infections and 46 thousand new deaths in the month of April^6^. Some of the currently authorized vaccines are mRNA-based and have unique cold storage and distribution requirements that present extra challenges to the world^17,18^. Vaccines that can offer improved safety and tolerability profiles could help overcome vaccine hesitancy. Furthermore, the development of vaccines that are temperature stable can help to improve the global distribution to countries with little or no capacity for cold storage^18^.

Most currently authorized SARS-CoV-2 vaccines specifically target the Spike glycoprotein which exists as a homotrimer on the virus surface^11,12,19^. The S1 domain of the protein contains a receptor binding-domain RBD that mediates attachment to the host cell angiotensin converting enzyme 2 (ACE2) receptor, while the S2 domain mediates entry through the host cell membrane^18^. Since all currently authorized vaccines contain both the S1 and S2 domains, inclusion of the full Spike protein alone in a vaccine appears to be reliably sufficient for driving an effective immune response.

Multiple clinical studies over the past decade have consistently shown a high level of safety and immunogenicity for plasmid DNA vaccines delivered into the cells by *in vivo* electroporation^2,3,20-22^. In addition to generating a humoral immune response, the DNA vaccine platform has been shown to consistently induce strong anti-viral T-cell responses that may not be elicited by other vaccine platforms, such as inactivated viruses^16,23^. Although much attention has been focused on the induction of anti-Spike protein antibodies, mounting evidence suggests that both humoral and cellular responses are required for prevention of COVID-19^24-26^. Robust T-cell responses are typically detected in convalescent donors despite many donors lacking measurable SARS-CoV-2 antibody^24,27,28^. Furthermore, a SARS-CoV-2 challenge study in non-human primates (NHPs) indicated that CD8 T-cells had a major contribution to the reduction in viral loads for animals with low anti-SARS-CoV-2 IgG levels^29^.

INO-4800 is plasmid DNA vaccine administered ID followed delivered by electroporation that encodes for both the S1 and S2 subunits of SARS-CoV-2 Spike protein^30^. It was previously shown in an open-label, Phase 1 study (NCT03721718)^31^ to have a favorable safety and tolerability profile, inducing humoral and or cellular responses in all 38 participants^31^. In that study, 1.0 mg or 2.0 mg doses of INO-4800 when administered ID followed by EP, lead to an increased immune response without added reactogenicity^31^. Additionally, a NHP challenge study demonstrated safety, immunogenicity, and efficacy of INO-4800 where vaccinated animals showed a robust anti-Spike humoral and T-cell response resulting in reduction in viral loads post-challenged^32^. We report the preliminary findings from the first randomized, blinded, placebo-controlled, Phase 2 clinical trial evaluating the safety and immunogenicity of INO-4800 in adults at high risk of SARS-CoV-2 exposure.

## Methods

### Study Design and Endpoints

The clinical trial is designed as a Phase 2/3, randomized, blinded, placebo-controlled, multi-center trial (NCT04642638) to evaluate the safety, immunogenicity, and efficacy of INO-4800 administered ID followed by electroporation (EP). The Phase 2 segment is designed to further evaluate the safety and immunogenicity of two doses of INO-4800 (1.0mg and 2.0mg) in a 2-dose regimen in SARS-CoV-2 seronegative adults to select the dose for efficacy evaluation in the Phase 3 segment. The findings reported here are applicable to the Phase 2 segment.

The clinical protocol was approved by a central and site-specific institutional review board. The conduct of the study was performed under current Good Clinical Practices. All participants provided written informed consent prior to enrollment. Healthy participants at least 18 years of age with a high risk of exposure to SARS-CoV-2 were randomized as described in Study Procedures section below. Participants were enrolled at 16 locations in the U.S.

The primary endpoints for the Phase 2 segment were immunologic in nature and comprised antigen-specific cellular immune responses measured by IFN-γ ELISpot assay and neutralizing antibody responses as measured by a pseudovirus-based neutralization assay. The secondary endpoints focused on safety and tolerability, measuring the incidence of solicited and unsolicited local and systemic reactions, including serious adverse events (SAEs) and adverse events of special interest (AESIs). For the purposes of this report, immunology endpoints were assessed at Week 6 (2 weeks post-dose 2) and safety and tolerability endpoints were assessed at Week 8. As specified in the clinical trial protocol, group-level unblinded interim summaries of the immunogenicity and safety data were produced, while maintaining subject-level blinding. Long-term follow-up data will continue to be collected for all subjects who have not discontinued with remaining visits through the final visit. Because subject-level blinding was to be maintained, not all of the adverse events can be displayed in by-treatment group summary tables.

### DNA Vaccine INO-4800

The vaccine was produced according to current Good Manufacturing Practices. INO-4800 contains plasmid pGX9501 expressing a synthetic, full-length sequence of the SARS-CoV-2 Spike glycoprotein of the original Wuhan strain, created using Inovio’s proprietary *in silico* Gene Optimization Algorithm to enhance expression as previously described^30^. The DNA sequence changes do not impact the amino acid sequence. The placebo group received equivalent volumes of saline sodium citrate buffer.

Electroporation following ID administration of INO-4800 is delivered using the CELLECTRA^®^ 2000 device that generates a controlled electric field at the injection site to enhance the cellular uptake and expression of the DNA plasmid. The device delivers a total of four electrical pulses per EP, each pulse of 52 msec in duration, at strengths of 0.2 Amp current and voltage of 40-200 V per pulse.

### Participant Eligibility

Eligible participants must have met the following criteria: healthy adults aged 18 years or older; able and willing to comply with all study procedures; individuals working or residing in an environment with high risk of exposure to SARS-CoV-2 for whom exposure may be relatively prolonged or for whom personal protective equipment may be inconsistently used; use of medically effective contraception with a failure rate of < 1% per year when used consistently, post-menopausal, or surgically sterile or have a partner who is sterile. Key exclusion criteria included the following: acute febrile illness with temperature higher than 100.4°F (38.0°C) or acute onset of upper or lower respiratory tract symptoms (e.g., cough, shortness of breath, sore throat); positive serologic or molecular (reverse transcription polymerase chain reaction [RT-PCR]) test for SARS-CoV-2 at Screening; pregnant or breastfeeding, or intending to become pregnant or intending to father children within the projected duration of the trial starting from the Screening visit until 3 months following the last dose; known history of uncontrolled HIV based on a CD4 count less than 200 cells/mm^3^ or a detectable viral load within the past 3 months; currently participating or has participated in a study with an investigational product within 30 days preceding Day 0; previous receipt of an investigational vaccine for prevention or treatment of COVID-19, MERS, or SARS (documented receipt of placebo in previous trial would be permissible for trial eligibility); respiratory diseases (e.g., asthma, chronic obstructive pulmonary disease) requiring significant changes in therapy or hospitalization for worsening disease during the 6 weeks prior to enrollment; immunosuppression as a result of underlying illness or treatment; lack of acceptable sites for ID injection and EP.

### Study Procedures

Eligible participants were randomized at a 3:3:1:1 ratio to receive one or two 1.0 mg ID injection(s) of INO-4800 or one or two ID injection(s) of placebo, followed by EP, administered at Days 0 and 28. The injection was administered in 0.1 mL volume over the deltoid or anterolateral quadriceps muscles followed by EP using CELLECTRA^®^ 2000 as previously described^31^. Participants in the 1.0 mg (or placebo) dose group received a single ID injection at each dosing visit with the second dose being administered similarly in a different limb (arm or leg) from the first dose. Participants in the 2.0 mg (or placebo) dose group received a single injection in 2 different limbs at each dosing visit.

Participants are assessed at screening, Days 0 (dose 1), 7, 28 (dose 2), 35, 42 (Week 6), 56 (Week 8), 210, and 392 post-dose 1. A participant diary was administered in the Phase 2 segment to collect solicited local and systemic AEs on the day of dosing and for 6 days following each dose. Local and systemic AEs, regardless of relationship to the vaccine, are assessed, recorded, and graded by the Investigator. Safety laboratory testing (complete blood count, comprehensive metabolic panel, and urinalysis) are collected at screening, Days 0, 28, 42, and 392 post-dose 1. AEs are graded according to the Common Terminology Criteria for Adverse Events (CTCAE) version 5.0. Injection site reactions are graded per the Toxicity Grading Scale for Healthy Adult and Adolescent Volunteers Enrolled in Preventive Vaccine Clinical Trials guidelines that were issued by the Food and Drug Administration in September 2007. An independent Data Safety Monitoring Board (DSMB) is chartered to review AE and laboratory data on a regular basis and reviewed the Day 56 (Week 8) safety data presented in this report. There are protocol-specified safety stopping rules.

If participants develop any symptoms suggestive of COVID-19, they are evaluated by the Investigator to include RT-PCR testing for SARS-CoV-2. Participants developing COVID-19 prior to receiving dose 2 were not permitted to receive dose 2.

Immunology specimens (cellular and humoral samples) were collected Days 0 (pre-dose), 42 (Week 6), 210, and 392 (post-dose 1).

### Immunogenicity Assessment Methods

For this report, samples collected at Day 0 (pre-dose) and Week 6 were analyzed. Peripheral blood mononuclear cells (PBMCs) were isolated from blood samples by a standard overlay on Ficoll-Hypaque followed by centrifugation. Isolated cells were frozen in 10% DMSO and 90% fetal calf serum. The frozen PBMCs were stored in liquid nitrogen for subsequent analyses. Serum samples were obtained fromwhole blood collection and stored at -80 °C until used to measure binding and neutralizing antibody titers.

#### SARS-CoV-2 Pseudovirus Neutralization Assay

SARS-CoV-2-DeltaCT pseudovirus was produced from HEK 293T cells transfected with GeneJammer (Agilent) using IgE-SARS-CoV-2 S plasmid (Genscript) and pNL4-3.Luc.R-E-plasmid (NIH AIDS reagent) at a 1:1 ratio. SARS-CoV-2-DeltaCT pseudovirus was titered to yield greater than 20 times the cells only control relative luminescence units (RLU) after 72h of infection. The assay was performed in a 96-well plate using 10,000 CHO cells stably expressing human ACE2 as target cells (Creative Biolabs, Catalog No. VCeL-Wyb019) in 100 µl D10 (DMEM supplemented with 10% FBS and 1X Penicillin-Streptomycin) media. On the following day, heat inactivated sera from INO-4800 vaccinated subjects were serially diluted as desired and incubated with a fixed amount of SARS-CoV-2-DeltaCT pseudovirus for 90 minutes at room temperature. The sera and pseudovirus mix were transferred to the plated cells and incubated for 72h. Cells were then lysed using britelite plus luminescence reporter gene assay system (Perkin Elmer Catalog no. 6066769) and RLU were measured using the Biotek plate reader. Neutralization titers (ID50) were defined as the reciprocal serum dilution at which RLU were reduced by 50% compared to RLU in virus control wells after subtraction of background RLU in cell control wells. Data for percent neutralization vs serum dilution was fitted to nonlinear regression i.e., log(inhibitor) vs. normalized response - Variable slope Least squares fit to obtain an ID_50_ value. All calculations were done using GraphPad Prism 8.

#### S1+S2 Enzyme-Linked Immunosorbent Assay (ELISA)

ELISA plates were coated with 2.0 µg/mL recombinant SARS-CoV-2 S1+S2 spike trimer protein (Acro Biosystems; SPN-C52H9) containing a C-terminal His tag, seven proline substitutions for trimer stabilization (F817P, A892P, A899P, A942P, K986P, V987P) and two mutations (R683A and R685A) to remove the furin cleavage sequence. The plates were then washed 4x with PBS with 0.05% Tween-20 (Sigma; P3563) and blocked (Starting Block, Thermo Scientific; 37538) for 1-3 hours. Serum samples were diluted a minimum of 1/20 in Starting Block and were added in duplicate to the washed and blocked assay plates. The samples were incubated for 2 hours at room temperature on a plate shaker set at 600 rpm. After washing three times in PBS containing 0.05% Tween-20, anti-human IgG HRP conjugate (BD Pharmingen; 555,788), diluted 1/1000 in Starting Block, was added to plates and incubated for 60 minutes at room temperature on a plate shaker set at 600 rpm. Plates were then washed three times in PBS containing 0.05% Tween-20, and TMB substrate (KPL; 5120-0077) was added to plates incubated for approximately nine minutes. Stop solution was added (KPL; 5150-0021), and optical density at 450 nm with background correction at 650 nm was read using a Synergy HTX Microplate Reader (BioTek). Antibody concentration in Units per mL (U/mL) were determined by interpolation from a four-parameter logistic model fit to a standard curve of reference convalescent plasma obtained >28 days after symptom onset from a PCR-confirmed SARS-CoV-2-recovered donor and arbitrarily assigned a concentration of 20,000 U/mL.

#### SARS-CoV-2 Spike ELISpot Assay Description

The ELISpot assay was performed as described previously^31^. Briefly, peripheral mononuclear cells (PBMCs) obtained at pre- and post-vaccination were stimulated overnight on precoated interferon-□ ELISpot plates (MabTech, Human IFN-□ ELISpot Plus) using overlapping 15-mer peptides comprising the entire spike protein sequence. Following overnight stimulation, ELISpot plates were processed for the detection of cellular IFN-□ production as according to the manufacturer’s instructions. Following the development of spots corresponding to cellular IFN-□ secretion, plates were scanned using a CTL S6 Micro Analyzer (CTL). Spots on the 96-well plates were counted using *ImmunoCapture* and *ImmunoSpot* software (CTL). Counts from negative control wells containing PBMCS with media only were subtracted from the counts of wells containing peptide stimulation. Reported values consisted of the mean counts across triplicate wells and were expressed as the number of spot forming units per million PBMCs.

#### Statistical Analysis

No formal power analysis was applicable to this trial. Descriptive statistics were used to summarize the safety endpoints: proportions with AEs, administration site reactions, and AESIs through 8 weeks. Descriptive statistics were also used to summarize the immunogenicity endpoints: post-baseline increases from baseline in interferon-□ ELISpot response magnitudes were compared between treatment groups using differences in medians and associated nonparametric 95% CIs for cellular results, and post-baseline increases from baseline in neutralizing antibody response titers were compared between treatment groups using ratios of geometric mean fold rises (GMFR) and associated t-distribution based 95% CIs.

## Results

### Study Population Demographics

Between November 30, 2020 and February 05, 2021, a total of 619 participants were screened, with a 35% screen-fail rate; and 401 participants were randomized by sixteen U.S. sites, each enrolling 6 to 55 participants. A total of 259 enrolled participants were 18-50 years of age. 142 were >51 years of age (32 were 65 years or older). A total of 201 participants were randomized to receive either INO-4800 as a 1.0 mg dose or placebo (both as single injections) and 200 participants were randomized to receive INO-4800 as a 2.0 mg dose or placebo (both as 2 injections per visit) with each injection followed by EP in a 2-dose regimen (Days 0 and 28) (Figure 1). Participants were 52.6% (211/401) female and were mostly white (84.0%, 337/401) with a median age of 44.4 years (range 18 to 80 years) (Table 1).

**Table 1:** Demographics by treatment groups

| Characteristics |  | Statistics | INO-4800<br>1.0 mg /Placebo<br>(N = 201) | INO-4800<br>2.0 mg/Placebo<br>(N = 200) | Total |
| --- | --- | --- | --- | --- | --- |
| **Race** |  |  |  |  |  |
| White |  | N (%) | 172 (85.6%) | 165 (82.5%) | 337 (84.0%) |
| Black |  | N (%) | 20 (10.0%) | 18 (9.0%) | 38 (9.5%) |
| American Indian |  | N (%) | 2 (1.0%) | 4 (2.0%) | 6 (1.5%) |
| Native Hawaiian |  | N (%) | 2 (1.0%) | 0 (0.0%) | 2 (0.5%) |
| Asian |  | N (%) | 3 (1.5%) | 10 (5.0%) | 13 (3.2%) |
| Other |  | N (%) | 2 (1.0%) | 3 (1.5%) | 5 (1.2%) |
| **Ethnicity** |  |  |  |  |  |
| Hispanic or Latino |  | N (%) | 36 (17.9%) | 23 (11.5%) | 59 (14.7%) |
| Not Hispanic or Latino |  | N (%) | 165 (82.1%) | 175 (87.5%) | 340 (84.8%) |
| **Sex** |  |  |  |  |  |
| Female |  | N (%) | 104 (51.7%) | 107 (53.5%) | 211 (52.6%) |
| Male |  | N (%) | 97 (48.3%) | 93 (46.5%) | 190 (47.4%) |
| **Age (years)** |  | mean (SD) | 44.0 (14.34) | 44.7 (13.68) | 44.4 (14.00) |
|  |  | median | 44 | 45 | 45 |
|  |  | min – max | 18 – 80 | 18 – 75 | 18 – 80 |
| 18-50 | Years old | N | 130 | 129 | 259 |
| ≥51 | Years old | N | 71 | 71 | 142 |
| ≥65* | Years old | N | 16 | 16 | 32 |

\*Subset of ≥51

**Figure 1:**
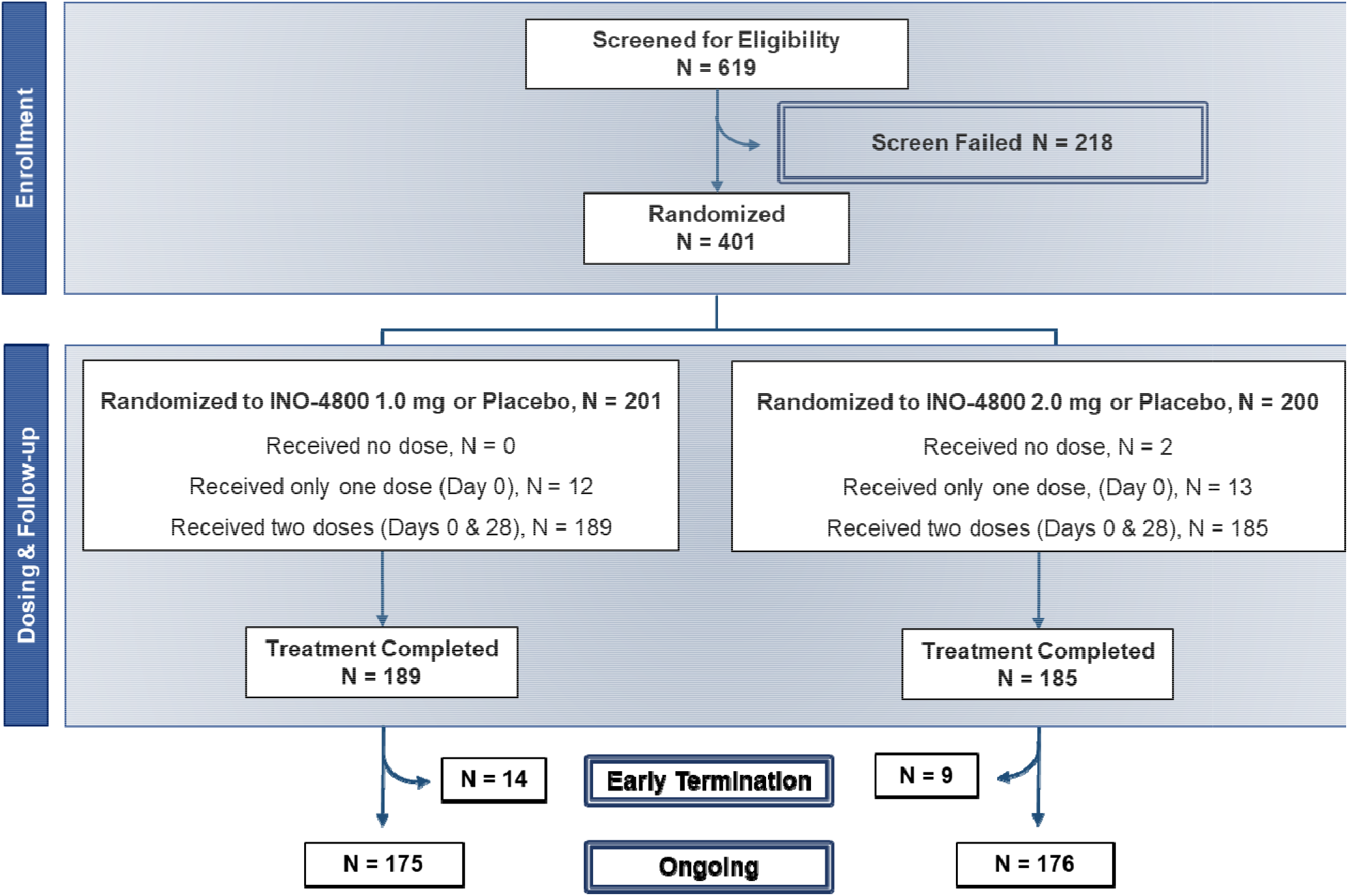
Phase 2 Enrollment Information

A total of 399 of 401 (99.5%) randomized participants were dosed and contributed to the safety population. Two randomized participants were not dosed due to loss to follow-up. Of the 399 participants who were dosed, 374 (93.3%) completed both doses. The reasons for not receiving the 2^nd^ dose were mainly due to opting to receive an emergency use authorized vaccine. A total of 374 participants completed a minimum follow-up of 28 days post-dose 2. A total of 23 participants were discontinued prior to Week 8 due to withdrawal by subject and lost to follow-up.

Of a total of 1153 ID injections administered to 399 participants, 1131 (98%) were administered in the arm and 22 in the leg (10 participants).

### Vaccine Safety and Tolerability

There was a total of 1,679 AEs recorded in 300 subjects through week 8. Of these, 1,446 treatment-related AEs were recorded in 281 subjects. The most common treatment-related (i.e., related to either investigational product or EP) AEs observed in greater than 5% of participants (Table 2) were injection site reactions (pruritus; 42 participants in 1.0 mg dose and 65 participants in 2.0 mg dose, pain; 35 participants in 1.0 mg dose and 41 participants in 2.0 mg dose, erythema; 25 participants in 1.0 mg dose and 36 participants in 2.0 mg dose, and swelling; 16 participants in 1.0 mg dose and 23 participants in 2.0 mg dose), fatigue (37 participants in 1.0 mg dose and 48 participants in 2.0 mg dose), headache (34 participants in 1.0 mg dose and 43 participants in 2.0 mg dose), myalgia/arthralgia (37 participants in 1.0 mg dose and 67 participants in 2.0 mg dose), and nausea (11 participants in 1.0 mg dose and 11 participants in 2.0 mg dose).

**Table 2:** Summary of treatment-related adverse events (>5% of Participants) by treatment

|  | **1.0 mg**<br>**INO-4800**<br>**(N = 151)** | **1 injection**<br>**Placebo**<br>**(N = 50)** | **2.0 mg**<br>**INO-4800**<br>**(N = 147)** | **2 injections**<br>**Placebo**<br>**(N = 51)** |
| --- | --- | --- | --- | --- |
|  | Participants<br>(%) | Participants<br>(%) | Participants<br>(%) | Participants<br>(%) |
| **Any AE\*** | 92 (60.9%) | 32 (64.0%) | 111 (75.5%) | 27 (52.9%) |
| **Local Reactions\*\*** |  |  |  |  |
| Injection site pruritus | 42 (27.8%) | 11 (22.0%) | 65 (44.2%) | 8 (15.7%) |
| Injection site pain | 35 (23.2%) | 8 (16.0%) | 41 (27.9%) | 9 (17.6%) |
| Injection site erythema | 25 (16.6%) | 10 (20.0%) | 36 (24.5%) | 4 (7.8%) |
| Injection site swelling | 16 (10.6%) | 1 (2.0%) | 23 (15.6%) | 1 (2.0%) |
| **Systemic Reactions** |  |  |  |  |
| Fatigue | 37 (24.5%) | 19 (38.0%) | 48 (32.7%) | 12 (23.5%) |
| Headache | 34 (22.5%) | 16 (32.0%) | 43 (29.3%) | 13 (25.5%) |
| Myalgia | 24 (15.9%) | 10 (20.0%) | 43 (29.3%) | 9 (17.6%) |
| Arthralgia | 13 (8.6%) | 4 (8.0%) | 24 (16.3%) | 5 (9.8%) |
| Nausea | 11 (7.3%) | 3 (6.0%) | 11 (7.5%) | 6 (11.8%) |

\*Primarily grade 1 and 2 AEs
\*\* Injection site bruising is not displayed due to ongoing blinded nature of the trial.
Note: Participants = unique number of subjects experiencing the adverse event. % = the proportion of subjects experiencing the event divided by the total in the safety population. MedDRA version 23.0 used for coding of Adverse Events.

The majority of AEs were Grade 1 and Grade 2 in severity and did not appear to increase in frequency with the second dose (Figures 2 and 3). Three Grade 3 AEs were reported: arthralgia (related to treatment), and cervical dysplasia and skin laceration (both not related to treatment). The single case of Grade 3 arthralgia occurred in a participant with a history of shoulder arthroscopy and in whom the arthralgia was limited to the previously injured shoulder. There were no Grade 4 AEs, no AESIs and no related SAEs. A single SAE of spontaneous abortion was assessed as not related to treatment. The number of participants experiencing each of the most common AEs did not differ appreciably between the two dosing groups (Table 2). The clinical plan is to follow the current Phase 2 participants for 12 months post-dose 2 for long-term safety.

**Figure 2:**
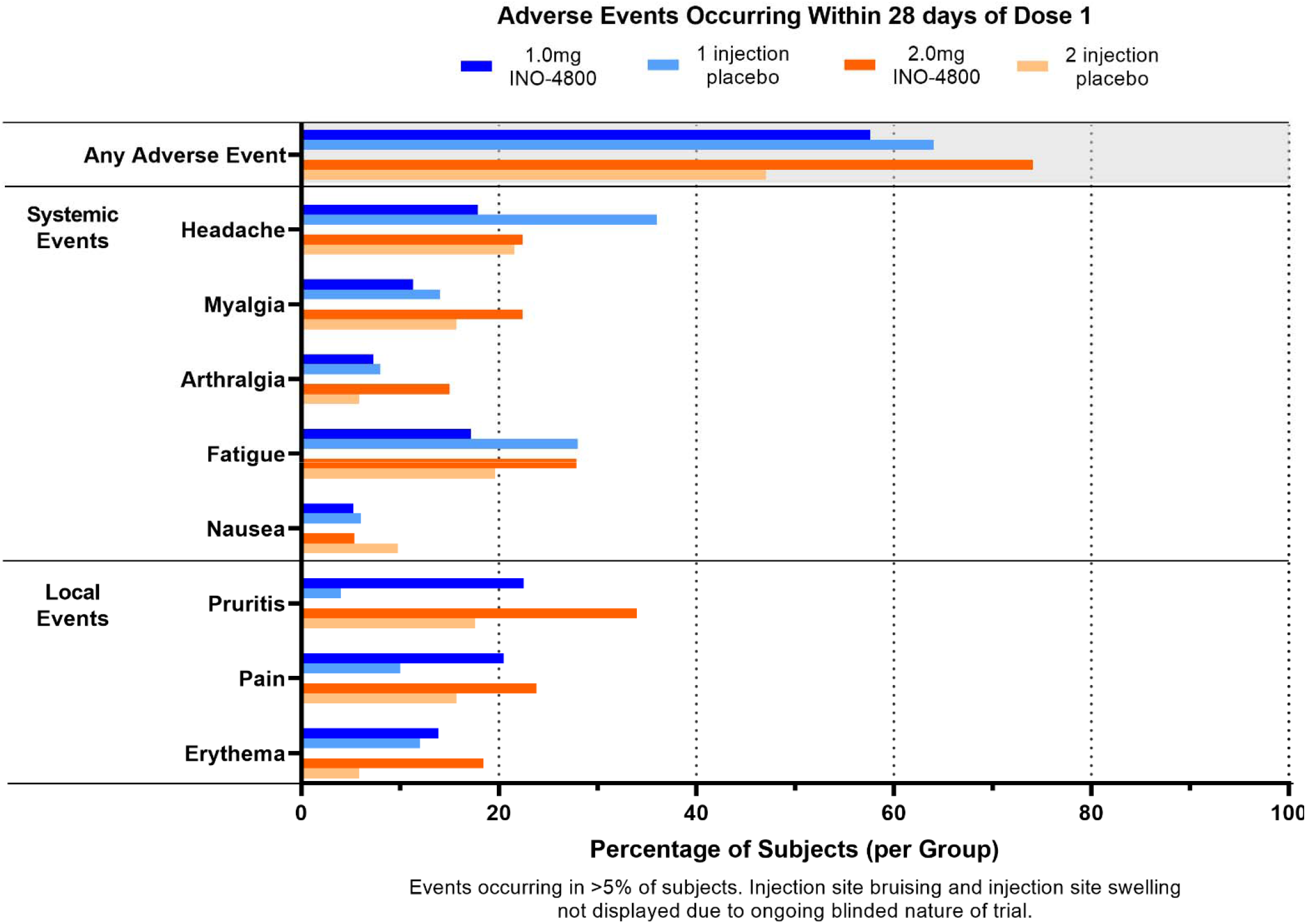
Adverse Events occurring within 28 days of dose 1

**Figure 3:**
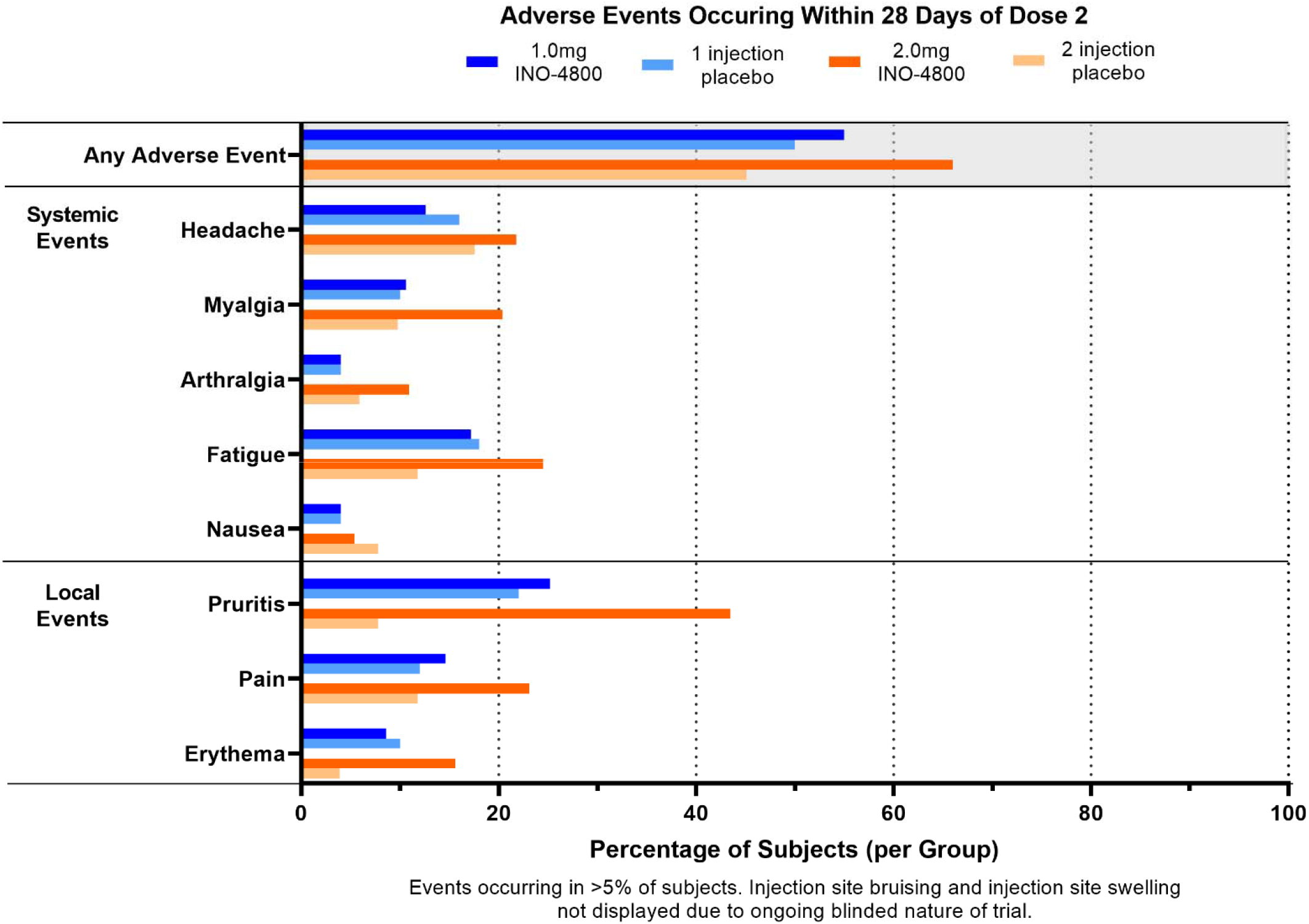
Adverse Events occurring within 28 days of dose 2

### Immunogenicity

Immunogenicity analyses included evaluating changes from baseline to Week 6 of binding antibody titers by ELISA, pseudovirus neutralizing antibody titers, and Interferon-□ ELISpot spot forming units (SFU). Subjects that completed two doses with Dose 2 at least 25 days after Dose 1, and who were not NP-positive were included in the analyses. Evaluable baseline samples and evaluable Week 6 samples that were at least 6 and at most 30 days after Dose 2 were included in the analyses.

### Humoral Immune Responses

Sera from placebo and INO-4800 participants were tested blindly for the ability to bind S1+S2 spike protein of SARS-CoV-2. At week 6, the geometric mean titers (GMT) (SD of log_10_) of binding antibody in the 1.0 mg and 2.0 mg dose groups were 938.8 (0.76) and 2210.0 (0.75) Units/ml (U/ml), respectively, compared with baseline GMT (SD) of 123.3 (0.68) and 93.5 (0.48) U/ml, respectively; the GMT (SD) of binding antibody in the 1- and 2-injection placebo groups at this timepoint were 92.8 (0.43) and 145.6 (0.54) U/ml, respectively, compared with baseline GMT (SD) of 110.2 (0.51) and 123.8 (0.52) U/ml, respectively (Table 3). The geometric mean fold rise (GMFR)(95%CI) was statistically significantly greater in the 1.0 mg dose group versus the 1 injection placebo group 8.34 (4.92, 14.14) as well as in the 2.0 mg dose group versus the 2 injection placebo group 19.99 (11.74, 34.03). The geometric mean fold rise (GMFR)(95%CI) was statistically significantly greater in the 2.0 mg dose group versus the 1.0 mg dose group 3.03 (2.04,4.44)

**Table 3:** Binding antibody responses by ELISA assay in all age groups

| Binding Antibody Titers |  | 1.0 mg<br>INO-4800 | 1 injection<br>Placebo | 2.0 mg<br>INO-4800 | 2 injections<br>Placebo |
| --- | --- | --- | --- | --- | --- |
| **Baseline** |  |  |  |  |  |
|  | GMT<br>(SD)<sup>a</sup> | 123.3<br>(0.68) | 110.2<br>(0.51) | 93.5<br>(0.48) | 123.8<br>(0.52) |
|  | N | 122 | 45 | 116 | 44 |
| **Week 6** |  |  |  |  |  |
|  | GMT<br>(SD)<sup>a</sup> | 938.8<br>(0.76) | 92.8<br>(0.43) | 2210.0<br>(0.75) | 145.6<br>(0.54) |
|  | N | 125 | 45 | 117 | 44 |
| **Change from baseline to**<br>**Week 6** |  |  |  |  |  |
|  | GMFR<br>(SD)<sup>a</sup> | 7.8<br>(0.74) | 0.9<br>(0.24) | 23.5<br>(0.78) | 1.2<br>(0.22) |
|  | N | 122 | 44 | 115 | 44 |

Abbreviation: GMT = Geometric Mean Titer, GMFR = Geometric Mean Fold Rise
Note: Baseline is defined as the last measurement prior to the first treatment administration.
GMT is calculated as $\text{anti-log}_{10}(\text{mean}[\log_{10} T_i])$ where $T_i$ is the assay result for subject $i$ . GMFR is calculated as $\text{anti-log}_{10}(\text{mean} [\log_{10} (Y_i/B_i)])$ where $Y_i$ is the post dose assay result for subject $i$ and $B_i$ is the baseline assay result for subject $i$ .
<sup>a</sup> Standard Deviation (SD) of the $\log_{10}$ titer values
GMT(SD) for COVID-19 convalescent donor plasma was 19444.3 (0.53).

Sera were also tested for the ability to neutralize SARS-CoV-2-DeltaCT pseudovirus. At week 6, the geometric mean titers (GMT) (SD of log_10_) of neutralizing antibody in the 1.0 mg and 2.0 mg dose groups were 93.6 (0.47) and 150.6 (0.46), respectively, compared with baseline GMT (SD) of 32.2 (0.38) and 35.8 (0.45), respectively (Table 4). The GMFR (SD) of neutralizing antibody at Week 6 relative to baseline in the 1.0 mg and 2.0 mg dose groups were 2.9 (0.45) and 4.3 (0.53), respectively; the GMFR (SD) of neutralizing antibody in the 1- and 2-injection placebo groups at this timepoint were 1.2 (0.32) and 1.0 (0.34), respectively (Table 4). The GMFR (95%CI) was statistically significantly greater in the 2.0 mg dose group versus the 1.0 mg dose group 1.47 (1.12,1.92).

**Table 4:** Neutralization antibody responses assessed by pseudotyped virus neutralization assay in all age groups

| Neutralizing Antibody Titers |  | 1.0 mg<br>INO-4800 | 1 injection<br>Placebo | 2.0 mg<br>INO-4800 | 2 injections<br>Placebo |
| --- | --- | --- | --- | --- | --- |
| **Baseline** |  |  |  |  |  |
|  | GMT<br>(SD)<sup>a</sup> | 32.2<br>(0.38) | 30.3<br>(0.40) | 35.8<br>(0.45) | 36.3<br>(0.43) |
|  | N | 124 | 46 | 114 | 43 |
| **Week 6** |  |  |  |  |  |
|  | GMT<br>(SD)<sup>a</sup> | 93.6<br>(0.47) | 32.5<br>(0.33) | 150.6<br>(0.46) | 35.3<br>(0.41) |
|  | N | 125 | 45 | 115 | 43 |
| **Change from baseline to**<br>**Week 6** |  |  |  |  |  |
|  | GMFR<br>(SD)<sup>a</sup> | 2.9<br>(0.45) | 1.2<br>(0.32) | 4.3<br>(0.53) | 1.0<br>(0.34) |
|  | N | 124 | 45 | 113 | 43 |

Abbreviation: GMT = Geometric Mean Titer, GMFR = Geometric Mean Fold Rise
Note: Baseline is defined as the last measurement prior to the first treatment administration.
GMT is calculated as $\text{anti-log}_{10}(\text{mean}[\log_{10} T_i])$ where $T_i$ is the assay result for subject $i$ . GMFR is calculated as $\text{anti-log}_{10}(\text{mean} [\log_{10} (Y_i/B_i)])$ where $Y_i$ is the post dose assay result for subject $i$ and $B_i$ is the baseline assay result for subject $i$ .
<sup>a</sup> Standard Deviation (SD) of the $\log_{10}$ titer values
GMT(SD) for COVID-19 convalescent donor plasma was 921.5 (0.51).

The binding antibody and neutralizing antibody responses were similar among different age groups. Supplementary tables are provided for binding antibody responses by ELISA in 18- to 50-year-olds (Table S1a), ≥ 51-year-olds (Table S1b), and ≥ 65-year-olds (Table S1c) and for pseudo neutralization data in 18- to 50-year-olds (Table S2a), ≥ 51-year-olds (Table S2b), and ≥ 65-year-olds (Table S2c).

### Enzyme-linked Immunospot (ELISpot)

Peripheral blood mononuclear cells (PBMCs) collected from study participants were tested blindly to measure T cell immune responses using the IFN-□ ELISpot assay. The median increase from baseline to Week 6 was 0.00 in both the 1- and 2-injection placebo groups, with max increases of 47.7 and 35.5 spot forming units (SFU) per 10^6^ PBMC observed in the 1- and 2-injection groups, respectively (Table 5). In the INO-4800 vaccinated groups, the median (min – max) increase from baseline to Week 6 was 3.40 (0.0 – 90.0) SFU per 10^6^ PBMC in the 1.0 mg dose group and 12.75 (0.0 – 465.0) SFU per 10^6^ PBMC in the 2.0 mg dose group (Table 5). Magnitudes of IFN-□ trended higher in the 1.0mg INO-4800 group compared to the 1-injection placebo group and were statistically significantly higher in the 2.0mg INO-4800 group compared to the 2-injection placebo group.

**Table 5:** T-cell immune responses by ELISpot assay in all age groups

| **Interferon-γ ELISpot**<br>**Spot-forming Units/$10^6$**<br>**PBMCs** |  | **1.0 mg**<br>**INO-4800** | **1 injection**<br>**Placebo** | **2.0 mg**<br>**INO-4800** | **2 injections**<br>**Placebo** |
| --- | --- | --- | --- | --- | --- |
| **Baseline** |  |  |  |  |  |
|  | median | 0.00 | 0.00 | 2.20 | 0.00 |
|  | min – max | 0.0 – 90.0 | 0.0 – 25.6 | 0.0 – 47.8 | 0.0 – 95.6 |
|  | N | 92 | 36 | 91 | 31 |
| **Week 6** |  |  |  |  |  |
|  | median | 6.70 | 3.30 | 18.90 | 2.20 |
|  | min – max | 0.0 – 96.7 | 0.0 – 63.3 | 0.0 – 468.3 | 0.0 – 131.1 |
|  | N | 108 | 40 | 97 | 37 |
| **Increase<sup>a</sup> from baseline**<br>**to Week 6** |  |  |  |  |  |
|  | median | 3.40 | 0.00 | 12.75 | 0.00 |
|  | min – max | 0.0 – 90.0 | 0.0 – 47.7 | 0.0 – 465.0 | 0.0 – 35.5 |
|  | N | 83 | 31 | 80 | 27 |

Note: Baseline is defined as the last measurement prior to the first treatment administration.
<sup>a</sup> If post-value is less than or equal to pre-value then increase=0.

T cell immune responses were similar across different age groups. Supplementary tables are provided for T-cell responses by ELISpot in 18- to 50-year-olds (Table S3a), ≥ 51-year-olds (Table S3b), and ≥ 65-year-olds (Table S3c).

## Discussion

We report initial data from the Phase 2 segment of an ongoing Phase 2/3 trial to evaluate the safety, tolerability, and immunogenicity of INO-4800, a SARS-CoV-2 vaccine encoding the spike protein (S), in adults at high risk of exposure to SARS-CoV-2. This 400-subject trial is intended to augment the Phase 1 trial data to select a dose for Phase 3 efficacy evaluation. The majority of AEs were Grade 1 and Grade 2 in severity, and importantly, did not appreciably appear to increase in frequency with the second dose. The only one case of treatment-related Grade 3 AE was arthralgia. A single SAE of spontaneous abortion was deemed not related to treatment.

INO-4800 generated balanced humoral and cellular immune responses in both 1.0 mg and 2.0 mg dose levels measured at Week 6 compared to the baseline levels at Day 0 (pre-dose) or compared to the placebo subjects at week 6 in all age groups tested. INO-4800 induced antibody responses in each INO-4800 dose group, which were capable of both binding and neutralization (Tables 3 and 4). At Week 6, both the 1.0 mg and 2.0 mg dose groups had GMFR values which were statistically significantly greater than each respective placebo group, and the 2.0 mg dose group was statistically higher than the 1.0 mg dose group.

The results were similar for the neutralizing antibody at Week 6, with the GMFR statistically significantly greater for both 1.0 mg and 2.0 mg dose groups versus the 1 injection and the 2 injection placebo groups, respectively. Not surprisingly, the GMFR of neutralizing antibody levels was statistically significantly greater in the 2.0 mg dose group versus the 1.0 mg dose group. The T cell immune responses measured by the ELISpot assay were also higher in the 2.0 mg dose group compared to the 1.0 mg dose group (Table 5). The T cell response results were similar to the previously published findings from Phase 1 trial, where INO-4800 vaccination led to substantial T cell responses with increased Th1 phenotype, measured by both IFN-□ ELISpot as well as multiparametric flow cytometry, as evidenced by increased expression of Th1-type cytokines IFN-□, TNF-α, and IL-2^31^. Assessment of cellular responses displayed the presence of SARS-CoV-2 specific CD4^+^ and CD8^+^ T cells exhibiting hallmarks of differentiation into both central and effector memory cells, suggesting that a persistent cellular response has been established^31^. Overall in Phase 2, the 2.0 mg dose group generated both binding and neutralizing antibody responses statistically significantly greater than those of 1.0 mg dose group while the T cell responses observed in the 2.0 mg dose group trended higher than those observed in the 1.0 mg dose group.

There are some interesting findings from this Phase 2 trial. Both binding and neutralizing antibody response levels generated with INO-4800 seemed higher in the Phase 2 trial compared to the same dose levels in Phase 1^31^. This might be explained by the increase in the sample size from Phase 1 to Phase 2. In addition, the consistency of a favorable tolerability profile from the first dose to the second dose in this Phase 2 safety data confirmed the Phase 1 finding of no increase in frequency of side effects after the second dose compared to the first dose. These results further suggest that in addition to being a primary vaccine candidate, INO-4800 could represent a safe booster vaccine without significant limitations such as anti-vector responses or dosing-incremented toxicities. We have observed directly in the Phase 1 trial participants who were boosted with a third dose of their INO-4800 vaccine between 6 and 10 months resulted in higher levels of humoral and cellular immune responses without increased levels of AEs (Manuscript in preparation). Given the uncertainty about the durability of the natural infection or vaccine induced responses against COVID-19 disease, vaccine boosting by a benign approach like INO-4800 may be an important way to maintain protection over subsequent epidemic waves of COVID-19. It is also possible that INO-4800 could serve as a useful booster shot for other S protein-targeted vaccine candidates with limitations in boosting ability, and this potential should be further investigated.

One of the major areas of research for COVID-19 is the investigation of the impact of recently emerged major variants of concern - first detected in the United Kingdom (B.1.1.7), South Africa (B.1.351), and Brazil (P.1) - on the protective efficacy of vaccines recently approved for emergency use as well as those vaccines currently in development. Previous studies indicated that neutralization activity in vaccinees’ sera only showed a minor reduction for B.1.1.7^34^, but neutralization was significantly decreased against B.1.351 and P.1^35-37^. That reduction of *in vitro* serum neutralization to B.1.351 seemed to correspond to a reduction of clinical vaccine efficacy^38-40^, while clinical efficacy to B.1.1.7 was more similar to wild type^34^. Similarly, reduced efficacy was found in countries such as Brazil that were suspected to have emerging P.1 variant infections during the trial^40^. We have previously tested and reported the humoral and cellular activity measured in Phase 1 trial INO-4800 vaccinated subjects against these SARS-CoV-2 variants. Similar to previous reports, there was a notable reduction in neutralization activity to B.1.351, while the reduction to B.1.1.7 was minor^36,41,42^. In contrast to studies for other vaccines, serum from INO-4800 vaccinees maintained neutralization activity to the P.1 variant as compared the wild type strain^35,43 42^. In addition, T cell responses generated by INO-4800 vaccination were consistently maintained between the wildtype and all SARS-CoV-2 variants tested, including B.1.351 and P.1^42^. The importance of T cell responses in preventing severe COVID-19 symptoms is supported by various studies of SARS-CoV-2 infection and convalescence even in the absence of antibodies^44-46^ and thus the ability to generate cross-reactive T cell responses of this type may prove critical in allowing for a single vaccine formulation to have coverage across multiple viral variants. These results further support the potential of INO-4800 protection in a global Phase 3 efficacy trial segment, which is currently being planned.

Furthermore, as current nucleic acid based COVID-19 vaccines roll out for administration in countries such as the United States, the requirement for a cold chain has posed significant logistical questions in regards to maintaining stability^17,33^. Conversely, INO-4800 has an excellent thermal stability profile, and based on current platform data is projected to be stable at room temperature for more than one year, at 37 degrees C for more than one month, and has a three to five-year projected shelf life at 2 to 8 degrees C (Unpublished data). Most significantly, INO-4800 does not require frozen cold-chain during transport or storage – a critical element when considering the feasibility of global distribution where the logistics of a cold chain become increasingly difficult.

Despite the early successes of vaccines approved for emergency use in many countries, the pandemic remains uncontrolled. The development of additional safe and effective vaccine platforms remains a global imperative. Vaccines, like INO-4800, if proven to be effective, could offer improved safety and tolerability as well as thermal stability that could be critical to controlling the pandemic in more remote areas where cold storage is unavailable or impractical. In addition to being a primary vaccine candidate, the potential for INO-4800 to be used as a booster is yet another potential benefit that supports the thesis to advance INO-4800 into Phase 3 efficacy evaluation to demonstrate effectiveness in the prevention of COVID-19.

## Supporting information

COVID19_311 Phase2 Supp. Material

## Data Availability

The preliminary COVID19-001 Phase 1 clinical study datasets are subject to access restriction to protect subject confidentiality as the clinical study is still ongoing.

## Contributors

MPM, PT and JA are co-first authors and drafted the manuscript. PT is the lead principal investigator for the trial. RS is the medical monitor for the trial.

JA, DA, ELR, MP, ACQ, NL, VMA, TRFS, SR, PP, AK, FIZ, DF, KL performed all clinical analytical assays.

JA, KK, AS, MPM, PP, performed and supervised all clinical analytical analyses. RJJ was responsible supervising for the cGMP production MD was responsible for statistical analyses.

MPM, KOS, HB, MG, EB, JC, GG, MCD, JL managed all activities related to the clinical study.

JDB, DBW, KEB, JES, LMH provided guidance for immune assays and data, and contributed to overall aspects of study related activities.

MPM, JL, MD, ASB, JES, JJK, KEB, JDB supervised all clinical and other study related activities.

## Acknowledgments

Inovio is grateful to all trial participants, the Investigators, Col Jessica Cowden (DoD/JPEO), and the Data Safety Monitoring Board for their important contributions to this research. We also recognize the outstanding and dedicated support of the clinical trial staff at 16 locations in the U.S.: Synexus Clinical Research U.S., Inc. – Phoenix Southeast in Chandler, Central Phoenix Synexus Clinical Research, and AMR Tempe (Arizona); Optimal Research, LLC in San Diego (California); AMR South Florida in Coral Gables and Clinical Research Trials of Florida, Inc. in Tampa (Florida); AMR Lexington (Kentucky); Walter Reed Army Institute of research in Silver Spring (Maryland); Ascension St. John Hospital in Detroit (Michigan), AMR Kansas City (Missouri); AMR, Clinical Research Consortium – Las Vegas (Nevada); University of Pennsylvania and Thomas Jefferson University in Philadelphia (Pennsylvania); Tekton Research in San Antonio and DM Clinical research in Tomball (Texas); and Advanced Clinical Research in West Jordan (Utah). We are also grateful to our Inovio team members who have steadfastly worked to combat the pandemic.

