## Supplementary material for "Safety and immunogenicity of INO-4800 DNA vaccine against SARS-CoV-2: a preliminary report of a randomized, blinded, placebo-controlled, Phase 2 clinical trial in adults at high risk of viral exposure": COVID19_311 Phase2 Supp. Material

**Supplemental Tables and Figures**

Table S1a: Binding antibody responses by ELISA in 18-to 50-year-olds

| **Binding Antibody Titers** |  | **1.0 mg** **INO-4800** | **1 injection** **Placebo** | **2.0 mg** **INO-4800** | **2 injections** **Placebo** |
| --- | --- | --- | --- | --- | --- |
| **Baseline** |  |  |  |  |  |
|  | GMT (SD)^a^ | 148.5 (0.76) | 135.1 (0.47) | 99.5 (0.41) | 118.7 (0.60) |
|  | N | 79 | 28 | 73 | 27 |
| **Week 6** |  |  |  |  |  |
|  | GMT (SD)^a^ | 1182.1 (0.79) | 122.9 (0.41) | 2671.2 (0.68) | 146.8 (0.66) |
|  | N | 80 | 27 | 73 | 27 |
| **Change from baseline to Week 6** |  |  |  |  |  |
|  | GMFR (SD)^a^ | 7.9 (0.71) | 1.0 (0.17) | 26.5 (0.73) | 1.2 (0.27) |
|  | N | 79 | 27 | 72 | 27 |

Abbreviation: GMT = Geometric Mean Titer, GMFR = Geometric Mean Fold Rise

Note: Baseline is defined as the last measurement prior to the first treatment administration.

GMT is calculated as anti-log_10_(mean[log_10_ Ti]) where Ti is the assay result for subject i. GMFR is calculated as anti-log_10_(mean [log_10_ (Yi/Bi)]) where Yi is the post dose assay result for subject i and Bi is the baseline assay result for subject i.

^a^ Standard Deviation (SD) of the log_10_ titer values

Table S1b: Binding antibody responses by ELISA in ≥51-year-olds

| **Binding Antibody Titers** |  | **1.0 mg** **INO-4800** | **1 injection** **Placebo** | **2.0 mg** **INO-4800** | **2 injections** **Placebo** |
| --- | --- | --- | --- | --- | --- |
| **Baseline** |  |  |  |  |  |
|  | GMT (SD)^a^ | 87.5 (0.48) | 78.9 (0.56) | 84.0 (0.59) | 132.4 (0.36) |
|  | N | 43 | 17 | 43 | 17 |
| **Week 6** |  |  |  |  |  |
|  | GMT (SD)^a^ | 623.3 (0.67) | 60.9 (0.41) | 1613.7 (0.85) | 143.7 (0.28) |
|  | N | 45 | 18 | 44 | 17 |
| **Change from baseline to Week 6** |  |  |  |  |  |
|  | GMFR (SD)^a^ | 7.6 (0.81) | 0.8 (0.33) | 19.2 (0.86) | 1.1 (0.12) |
|  | N | 43 | 17 | 43 | 17 |

Abbreviation: GMT = Geometric Mean Titer, GMFR = Geometric Mean Fold Rise

Note: Baseline is defined as the last measurement prior to the first treatment administration.

GMT is calculated as anti-log_10_(mean[log_10_ Ti]) where Ti is the assay result for subject i. GMFR is calculated as anti-log_10_(mean [log_10_ (Yi/Bi)]) where Yi is the post dose assay result for subject i and Bi is the baseline assay result for subject i.

^a^ Standard Deviation (SD) of the log_10_ titer values

Table S1c: Binding antibody responses by ELISA in ≥65-year-olds

| **Binding Antibody Titers** |  | **1.0 mg** **INO-4800** | **1 injection** **Placebo** | **2.0 mg** **INO-4800** | **2 injections** **Placebo** |
| --- | --- | --- | --- | --- | --- |
| **Baseline** |  |  |  |  |  |
|  | GMT (SD)^a^ | 63.8 (0.40) | 48.4 (0.38) | 55.1 (0.39) | 141.6 (0.29) |
|  | N | 9 | 4 | 12 | 4 |
| **Week 6** |  |  |  |  |  |
|  | GMT (SD)^a^ | 538.3 (1.01) | 31.3 (0.00) | 1216.0 (1.19) | 142.9 (0.20) |
|  | N | 10 | 4 | 12 | 4 |
| **Change from baseline to Week 6** |  |  |  |  |  |
|  | GMFR (SD)^a^ | 11.6 (1.09) | 0.6 (0.38) | 22.1 (1.12) | 1.0 (0.10) |
|  | N | 9 | 4 | 12 | 4 |

Abbreviation: GMT = Geometric Mean Titer, GMFR = Geometric Mean Fold Rise

Note: Baseline is defined as the last measurement prior to the first treatment administration. GMT is calculated as anti-log_10_(mean[log_10_ Ti]) where Ti is the assay result for subject i. GMFR is calculated as anti-log_10_(mean [log_10_ (Yi/Bi)]) where Yi is the post dose assay result for subject i and Bi is the baseline assay result for subject i.

^a^ Standard Deviation (SD) of the log_10_ titer values

Table S2a: Neutralization antibody responses assessed by pseudotyped virus neutralization assay in 18-to 50-year-olds

| **Neutralizing Antibody Titers** |  | **1.0 mg** **INO-4800** | **1 injection** **Placebo** | **2.0 mg** **INO-4800** | **2 injections** **Placebo** |
| --- | --- | --- | --- | --- | --- |
| **Baseline** |  |  |  |  |  |
|  | GMT (SD)^a^ | 34.5 (0.41) | 34.4 (0.44) | 32.2 (0.42) | 38.3 (0.47) |
|  | N | 80 | 28 | 72 | 27 |
| **Week 6** |  |  |  |  |  |
|  | GMT (SD)^a^ | 112.6 (0.49) | 33.5 (0.34) | 159.9 (0.43) | 42.3 (0.44) |
|  | N | 80 | 27 | 73 | 27 |
| **Change from baseline to Week 6** |  |  |  |  |  |
|  | GMFR (SD)^a^ | 3.3 (0.47) | 1.1 (0.33) | 5.0 (0.57) | 1.1 (0.41) |
|  | N | 80 | 27 | 72 | 27 |

Abbreviation: GMT = Geometric Mean Titer, GMFR = Geometric Mean Fold Rise

Note: Baseline is defined as the last measurement prior to the first treatment administration. GMT is calculated as anti-log_10_(mean[log_10_ Ti]) where Ti is the assay result for subject i. GMFR is calculated as anti-log_10_(mean [log_10_ (Yi/Bi)]) where Yi is the post dose assay result for subject i and Bi is the baseline assay result for subject i.

^a^ Standard Deviation (SD) of the log_10_ titer values

Table S2b: Neutralization antibody responses assessed by pseudotyped virus neutralization assay in ≥51-year-olds

| **Neutralizing Antibody Titers** |  | **1.0 mg** **INO-4800** | **1 injection** **Placebo** | **2.0 mg** **INO-4800** | **2 injections** **Placebo** |
| --- | --- | --- | --- | --- | --- |
| **Baseline** |  |  |  |  |  |
|  | GMT (SD)^a^ | 28.4 (0.30) | 24.9 (0.32) | 43.0 (0.48) | 33.3 (0.34) |
|  | N | 44 | 18 | 42 | 16 |
| **Week 6** |  |  |  |  |  |
|  | GMT (SD)^a^ | 67.4 (0.40) | 31.2 (0.32) | 135.7 (0.51) | 26.1 (0.32) |
|  | N | 45 | 18 | 42 | 16 |
| **Change from baseline to Week 6** |  |  |  |  |  |
|  | GMFR (SD)^a^ | 2.3 (0.40) | 1.3 (0.30) | 3.2 (0.43) | 0.8 (0.16) |
|  | N | 44 | 18 | 41 | 16 |

Abbreviation: GMT = Geometric Mean Titer, GMFR = Geometric Mean Fold Rise

Note: Baseline is defined as the last measurement prior to the first treatment administration. GMT is calculated as anti-log_10_(mean[log_10_ Ti]) where Ti is the assay result for subject i. GMFR is calculated as anti-log_10_(mean [log_10_ (Yi/Bi)]) where Yi is the post dose assay result for subject i and Bi is the baseline assay result for subject i.

^a^ Standard Deviation (SD) of the log_10_ titer values

Table S2c: Neutralization antibody responses assessed by pseudotyped virus neutralization assay in ≥65-year-olds

| **Neutralizing Antibody Titers** |  | **1.0 mg** **INO-4800** | **1 injection** **Placebo** | **2.0 mg** **INO-4800** | **2 injections** **Placebo** |
| --- | --- | --- | --- | --- | --- |
| **Baseline** |  |  |  |  |  |
|  | GMT (SD)^a^ | 24.0 (0.34) | 20.9 (0.40) | 35.2  (0.32) | 35.3 (0.07) |
|  | N | 10 | 4 | 11 | 3 |
| **Week 6** |  |  |  |  |  |
|  | GMT (SD)^a^ | 76.9 (0.55) | 28.3 (0.34) | 125.3 (0.55) | 32.6 (0.12) |
|  | N | 10 | 4 | 11 | 3 |
| **Change from baseline to Week 6** |  |  |  |  |  |
|  | GMFR (SD)^a^ | 3.2 (0.45) | 1.4 (0.35) | 3.6 (0.58) | 0.9 (0.05) |
|  | N | 10 | 4 | 11 | 3 |

Abbreviation: GMT = Geometric Mean Titer, GMFR = Geometric Mean Fold Rise

Note: Baseline is defined as the last measurement prior to the first treatment administration. GMT is calculated as anti-log_10_(mean[log_10_ Ti]) where Ti is the assay result for subject i. GMFR is calculated as anti-log_10_(mean [log_10_ (Yi/Bi)]) where

Yi is the post dose assay result for subject i and Bi is the baseline assay result for subject i.

^a^ Standard Deviation (SD) of the log_10_ titer values

Table S3a: T-cell immune responses by ELISpot assay in 18-to 50-year-olds

| **Interferon-ɣ ELISpot Spot-forming Units** |  | **1.0 mg** **INO-4800** | **1 injection** **Placebo** | **2.0 mg** **INO-4800** | **2 injections** **Placebo** |
| --- | --- | --- | --- | --- | --- |
| **Baseline** |  |  |  |  |  |
|  | median | 1.10 | 0.00 | 0.55 | 0.00 |
|  | min – max | 0.0 – 90.0 | 0.0 – 25.6 | 0.0 – 47.8 | 0.0 – 47.8 |
|  | N | 55 | 19 | 56 | 18 |
| **Week 6** |  |  |  |  |  |
|  | median | 6.70 | 3.30 | 18.90 | 0.00 |
|  | min – max | 0.0 – 96.7 | 0.0 – 60.0 | 0.0 – 311.1 | 0.0 – 20.0 |
|  | N | 71 | 25 | 59 | 24 |
| **Increase^a^ from baseline to Week 6** |  |  |  |  |  |
|  | median | 3.30 | 0.00 | 12.20 | 0.00 |
|  | min – max | 0.0 – 90.0 | 0.0 – 16.7 | 0.0 – 311.1 | 0.0 – 15.6 |
|  | N | 50 | 17 | 49 | 16 |

Note: Baseline is defined as the last measurement prior to the first treatment administration.

^a^ If post-value is less than or equal to pre-value then increase=0.

Table S3b: T-cell immune responses by ELISpot assay in ≥51-year-olds

| **Interferon-ɣ ELISpot Spot-forming Units** |  | **1.0 mg** **INO-4800** | **1 injection** **Placebo** **(N=18)** | **2.0 mg** **INO-4800** **(N=42)** | **2 injections** **Placebo** **(N=16)** |
| --- | --- | --- | --- | --- | --- |
| **Baseline** |  |  |  |  |  |
|  | median | 0.00 | 0.00 | 3.30 | 1.10 |
|  | min – max | 0.0 – 42.2 | 0.0 – 16.7 | 0.0 – 30.0 | 0.0 – 95.6 |
|  | N | 37 | 17 | 35 | 13 |
| **Week 6** |  |  |  |  |  |
|  | median | 10.00 | 3.30 | 18.35 | 4.40 |
|  | min – max | 0.0 – 64.4 | 0.0 – 63.3 | 0.0 – 468.3 | 0.0 – 131.1 |
|  | N | 37 | 15 | 38 | 13 |
| **Increase^a^ from baseline to Week 6** |  |  |  |  |  |
|  | median | 6.70 | 0.55 | 13.30 | 4.40 |
|  | min – max | 0.0 – 64.4 | 0.0 – 47.7 | 0.0 – 465.0 | 0.0 – 35.5 |
|  | N | 33 | 14 | 31 | 11 |

Note: Baseline is defined as the last measurement prior to the first treatment administration.

^a^ If post-value is less than or equal to pre-value then increase=0.

Table S3c: T-cell immune responses by ELISpot assay in ≥65-year-olds

| **Interferon-ɣ ELISpot Spot-forming Units** |  | **1.0 mg** **INO-4800** | **1 injection** **Placebo** | **2.0 mg** **INO-4800** | **2 injections** **Placebo** |
| --- | --- | --- | --- | --- | --- |
| **Baseline** |  |  |  |  |  |
|  | median | 0.00 | 1.10 | 3.30 | 2.80 |
|  | min – max | 0.0 – 7.8 | 0.0 – 16.7 | 0.0 – 16.7 | 0.0 – 12.2 |
|  | N | 7 | 4 | 10 | 4 |
| **Week 6** |  |  |  |  |  |
|  | median | 5.60 | 1.65 | 13.30 | 3.85 |
|  | min – max | 0.0 – 24.4 | 0.0 – 3.3 | 0.0 – 468.3 | 3.3 – 4.4 |
|  | N | 7 | 2 | 11 | 2 |
| **Increase^a^ from baseline to Week 6** |  |  |  |  |  |
|  | median | 6.70 | 0.55 | 22.20 | 2.20 |
|  | min – max | 0.0 – 18.8 | 0.0 – 1.1 | 0.0 – 465.0 | 0.0 – 4.4 |
|  | N | 6 | 2 | 10 | 2 |

Note: Baseline is defined as the last measurement prior to the first treatment administration.

^a^ If post-value is less than or equal to pre-value then increase=0.
